# Impact of a natural disaster on access to care and biopsychosocial outcomes among Hispanic/Latino cancer survivors

**DOI:** 10.1101/19009704

**Authors:** Mary Rodriguez-Rabassa, Ruthmarie Hernandez, Zindie Rodriguez, Claudia B. Colon-Echevarria, Lizette Maldonado, Nelmit Tollinchi, Estefania Torres, Adnil Mulero, Daniela Albors, Jaileene Perez-Morales, Idhaliz Flores, Heather Jim, Eida M. Castro, Guillermo N. Armaiz-Pena

**Affiliations:** Clinical Psychology Program, School of Behavior and Brain Sciences, Ponce Health Sciences University, Ponce, Puerto Rico; Division of Mental Health, Ponce Research Institute, Ponce, Puerto Rico; Division of Cancer Biology, Ponce Research Institute, Ponce, Puerto Rico; Division of Women’s Health, Ponce Research Institute, Ponce, Puerto Rico; Department of Basic Sciences, Division of Pharmacology and Ponce Health Sciences University, Ponce, Puerto Rico; Department of Basic Sciences, Division of Microbiology, School of Medicine, Ponce Health Sciences University, Ponce, Puerto Rico; Department of Cancer Epidemiology, H. Lee Moffitt Cancer Center and Research Institute, Tampa, Florida; Department of Health Outcomes and Behavior, H. Lee Moffitt Cancer Center and Research Institute, Tampa, Florida

**Keywords:** biopsychosocial, cancer, natural disasters, access to care, cytokines

## Abstract

Cancer is the leading cause of death in Puerto Rico (PR). Hurricane Maria (HM) and its aftermath lead to widespread devastation in the island, including the collapse of the healthcare system. Medically fragile populations, such as cancer survivors, were significantly affected. The goal of this study was to assess the impact of HM on barriers to care, emotional distress, and inflammatory biomarkers among cancer survivors in PR. This exploratory longitudinal study was conducted in health care facilities and community support groups from PR. Cancer survivors (n=50) and non-cancer participants (n=50) completed psychosocial questionnaires and provided blood samples that were used to assess inflammatory cytokines levels. Data were analyzed through descriptive, frequencies, correlational, and linear regression analyses. Cancer survivors that were affected by HM reported increased barriers in accessing medical care, which were directly associated with anxiety, perceived stress, and post-traumatic symptomatology. Moreover, being a cancer survivor, along with closeness in time from HM predicted more barriers to receiving health care. Several inflammatory cytokines, such as CD31, BDNF, TFF3, Serpin E-1, Vitamin D BP, VCAM-1, Osteopontin, Chitinase 3 like 1, MMP-9 and MIF were significantly upregulated in cancer survivors while BDNF, MMP9 and Osteopontin had significant positive correlations with barriers to care. HM significantly impacted Puerto Ricans psychosocial well-being. Cancer survivors had significant barriers to care and showed increased serum inflammatory cytokines, but didn’t show differences in anxiety, stress and post-traumatic symptoms compared to non-cancer participants.

## Introduction

Natural disasters can significantly alter an individual’s daily life and lead to psychological distress, particularly in medically fragile populations such as cancer survivors. Hurricane Maria made landfall in Puerto Rico (PR) on September 20, 2017 as Category 4 storm, killing an estimated 2,975 people and causing an estimated $90 million in damages (1-3). Widespread devastation included loss of power and potable water infrastructure; destruction of buildings, bridges, and roads; lack of telecommunications; and closing of ports and airports (4). Lack of access to food and clean water was a significant problem for residents of PR (4). Mudslides rendered many roads in rural areas impassable, limiting relief efforts and access to medical care (4). The severity and duration of the aftermath caused significant psychological distress in the Puerto Rican population (5). Suicide rates rose by 29% compared to the year before (2016) (5). Moreover, during the subsequent three months after HM made landfall, the government psychosocial helpline received 2.4 times more calls to manage suicidal attempts (5). However, knowledge about psychological distress and biobehavioral factors after a natural disaster, such as HM, in cancer patients remains limited.

There is growing evidence supporting the role of stress in cancer progression (6). Several studies have suggested that altered psychological states, including chronic stress or depression, may accelerate growth of existing tumors (7). Planning for short- and long-term patient cancer care requires a holistic approach that considers the impact of psychological distress at the biological level in the context of a natural disaster. For example, traumatic stress, anxiety, and depression are common and persistent after a positive cancer diagnosis, and have been associated to alterations in circulating markers of inflammation (e.g., decreased NK cell cytotoxicity, elevated white blood cell counts, increased inflammatory macrophages) (7, 8). Moreover, extreme environmental stressors could have a significant effect on cancer survivors and lead to increased prevalence of psychological distress, such as, chronic stress and depression, that have been associated to disease progression (6). At the physiological level, activation of the sympathetic nervous system (SNS) or the hypothalamic-pituitary adrenal (HPA) axis lead to the release of norepinephrine, epinephrine, and cortisol, respectively. These have been shown to induce the release of immune and inflammatory factors (6). Specifically, activation of SNS and HPA resulted in elevated levels of several cytokines, infiltration of macrophages, and enhanced tumor growth (8, 9). In light of these data, it is important to determine whether and to what extent psychological distress (caused by an extreme environmental stressor) can contribute to altered behavioral states and systemic cytokine levels.

## Materials and Methods

The study followed the ethical principles from the Declaration of Helsinki and was approved by the Institutional Review Board of the Ponce Medical School Foundation. (IRB Approval 080121-IF). We obtained written informed consent from all participants at the time of enrollment.

### Participants

This prospective longitudinal case-control study collected participants’ blood and self-reported measurements. All measurements were obtained at the time of recruitment and every three months for a period of one year following the landfall of Hurricane Maria (September 20, 2017). Recruitment started four months after HM (January 2017) and up to August 2019 is still ongoing. Recruitment took place in health care facilities located in the southern area of PR, with the collaboration of community support groups and the PRBB facilities at a tertiary hospital in Ponce, PR. All participants were in the island at the time of HM and remained in PR for at least three months after the passage of the hurricane. The inclusion criteria were: participants must be between the ages of 21 and 89, with or without current or past history of any cancer type. Subjects were excluded if they self-reported or documented severe or uncontrolled psychiatric or neurological conditions that preclude their participation in the study.

### Natural Disaster Outcomes

The research team developed the Natural Disaster Outcomes Questionnaire, based on their personal experiences with HM, to identify problems suffered by participants in the aftermath of the hurricane. This survey contains 23 Likert-type questions that explores if and to what degree participants were impacted by hurricane-related problems during the previous three. Total scores range from 0 to 92, with higher scores reflecting higher impact. The questionnaire is included as Supplementary Data 1. This questionnaire was subjected to content validation by individuals that experienced HM and its aftermath.

### Psychological Distress and Access to Health Care

Participants answered a battery of questionnaires that explored psychological distress. Scoring and interpretation of these questionnaires followed the guidelines and procedures reported in the literature. To gather participants’ reports of depression and anxiety symptomatology they answered the PHQ-8 and the GAD-7 questionnaires, respectively (10-12). Participants were asked to complete the Distress Thermometer (psychological discomfort), the PSS (stressful situations), the PCL-5 (PTSD symptomatology) (13-15), BRS (resilience) and the PTGI-SF (post-traumatic growth) (16-18). Perception of social support was also assessed using the ISEL-12 (19). To assess potential barriers that participants faced with access to health care after the hurricane we utilized the BCQ (20, 21). To better understand the study results, the research team inverted the scoring system so higher scores would now indicate higher barriers.

#### Blood processing, storage and cytokine array analyses

Blood samples were obtained at the time of recruitment and processed within four hours of collection time to isolate serum following standard methods. Serum samples were stored at −80°C. Serum was analyzed with R & D Proteome Profiler Human XL Cytokine Array kits (Minneapolis, MN) according to manufacturer’s instructions. Arrays were quantified using Quick Spots Tool in Western Vision’s HLImage++ (Version 22.0). Cytokine heatmaps were constructed in RStudio (Version 1.0.153) with the following R packages: RColorBrewer, d3heatmap and ggplot2. To identify significant differences in cytokine patterns between cancer patients and controls, volcano plots were constructed using MultiExperiment Viewer Software (version 10.2). Protein-protein interaction network and gene enrichment of differentially expressed cytokines were constructed with STRING online platform (version 11.0).

### Statistical Analysis

Variable distributions were assessed with the Kolmogorov-Smirnov test to determine the statistical approach required. With the exception of the PSS Scale, scores for all variables were not normally distributed. We used non-parametric methods to describe the outcomes (e.g., median and interquartile ranks) based on participants’ history of cancer. We used Spearman Rho correlations to explore associations between variables and Mann-Whitney U tests to test group differences for the outcomes including cytokine levels. We performed linear regression analyses to predict barriers in access to care by cancer status, age, and time of recruitment after HM made landfall. We used the RStudio statistical program with the following packages: nortest, psych, olsrr, qgraph, dplyr, Hmisc, corrplot, tidyselect, ggpubr, gplots, RColorBrewer, d3heatmap and ggplot2. All tests were 2-sided and statistical significance was defined as *p*<0.05. Cytokine expression analysis were performed using expression raw values (arbitrary units) and differences in cytokine values between cancer and non-cancer. Cytokine distribution normality was assessed using the Shapiro-Wilk test. We performed Mann-Whitney analyses to identify significant differences in cytokine expression between groups. Statistical significance threshold for cytokine analyses was established at p<0.01.

## Results

### Study Population

Here we studied the biopsychosocial effect of a natural disaster (HM) on 100 Hispanic/Latino participants (50 cancer survivors and 50 non-cancer participants). The mean time of recruitment after the hurricane was 7.97 months (range: 4.33 – 11.27 months). Table 1 shows sociodemographic characteristics of study participants and data obtained from psychosocial questionnaires administered to all participants. In general, the majority of the participants were women (72%). Breast (44%) and prostate (14%) cancers were the most prevalent cancer types among cancer survivors. The age of cancer survivors [median (Mdn) = 58.5 (interquartile range (IQR) = 50.5-66.0)] was significantly higher from non-cancer participants (Mdn=49.5, IQR=38.5-57.0), *p*<0.05.

**Table 1.**
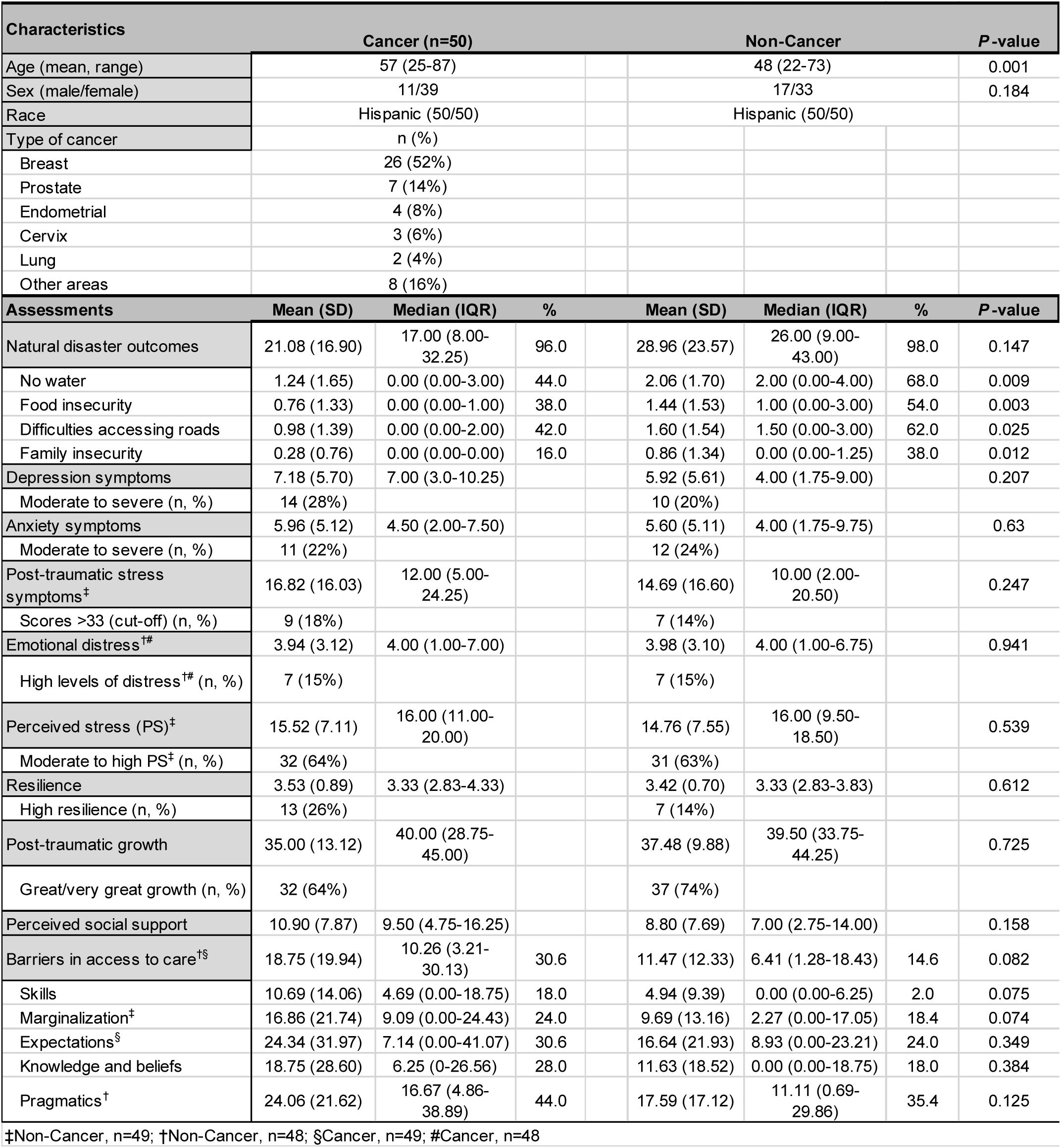
Clinical, demographical and psychosocial measurements from study participants.

### Natural Disaster Outcomes

Participants reported being negatively affected by all items identified in the Natural Disaster Outcomes Questionnaire (Table 1). Cancer survivors showed lower median total scores (Mdn=17.00, IQR=8.00-32.25) than non-cancer participants (Mdn=26.00, IQR=9.00-43.00) but this difference was not significant (*p*=0.146). The most frequent problems faced by participants from both groups were no electricity service (cancer: 68%; non-cancer: 86%) and difficulties with communication services (cancer: 66%; non-cancer: 82%), but these rates were not significantly different between groups. However, reports on the degree of discomfort caused by natural disaster related outcomes were usually higher in non-cancer participants than cancer survivors. For example, non-cancer participants reported higher rates of being affected by having no water service, food insecurity, difficulties accessing roads, and loss of security for the family (*p*<0.05).

### Psychological Distress

Cancer survivors showed higher symptomatology in several measures of psychological distress compared to non-cancer participants (Table 1). The median scores of depression symptomatology for cancer survivors were 7.00 (IQR=3.0-10.25) versus 4.00 (IQR=1.75-9.00) in non-cancer participants *(p*=0.21). Analyses of the severity of symptoms showed that a higher number of cancer survivors (n=14, 28%) are distributed in the categories of moderate to severe depression compared to non-cancer participants (n=10, 20%). Similarly, post-traumatic symptomatology scores in cancer survivors were slightly higher than non-cancer participants (Mdn=12.00, IQR=5.00-24.25 vs. Mdn=10.00, IQR=2.00-20.50; *p*=0.25). Moreover, increased severity of post-traumatic symptomatology (scores higher than 33) were more frequently observed in cancer survivors (cancer 18%, non-cancer 14%), who reported higher difficulties sleeping and concentrating or feeling easily startled.

Both groups revealed comparable scores on measures of anxiety symptomatology that were not significantly different (cancer: Mdn=4.50, IQR=2.00-7.50; non-cancer: Mdn=4.00, IQR=1.75-9.75), emotional distress (cancer: Mdn=4.00, IQR=1.00-7.00; non-cancer: Mdn=4.00, IQR=1.00-6.75), and perceived stress (cancer: Mdn=16.00, IQR=11.00-20.00; non-cancer: Mdn=16.00, IQR=9.50-18.50). Scores on resilience (cancer: Mdn=3.33, IQR=2.83-4.33; non-cancer: Mdn=3.33, IQR=2.83-3.83), post-traumatic growth (cancer: Mdn=40.00, IQR=28.75-45.00; non-cancer: Mdn=39.50, IQR=33.75-44.25) and perception in social support (cancer: Mdn=9.50, IQR=4.75-16.25; non-cancer: Mdn=7.00, IQR=2.75-14.00) were also not significantly different.

### Access to Health Care

Cancer survivors reported higher barriers in access to care compared to non-cancer participants (Table 1). Cancer survivors had higher median scores on the total scale (cancer: Mdn=10.26, IQR=3.21-30.13 vs non-cancer: Mdn=6.41, IQR=1.28-18.43) as well as in the Skills subscale (cancer: Mdn=4.69, IQR=0.00-18.75 vs. non-cancer: Mdn=0.00, IQR=0.00-6.25), but these differences did not reach statistical significance (*p*=0.08). The median score on the Marginalization subscale was greater in cancer survivors compared to non-cancer participants (Mdn=9.00, IQR=0.00-24.43 vs. Mdn=2.27, IQR=0.00-17.05, respectively) although not significant (*p*=0.07).

Next, we explored the associations between participants’ scores on various psychosocial questionnaires used in this study for each group, using the Spearman’s Rho correlation test. Figure 1 is a visual representation of a correlation network with significant associations (*p*<0.05) in cancer (A) and non-cancer participants (B), generated using qgraph in RStudio 1.1.463. Both groups displayed a direct correlation between scores of depression, anxiety, post-traumatic symptoms, distress, and perceived stress. Resilience scores were inversely related to these measures in all participants, except for depression, which showed no association. Resilience was also negatively associated with barriers to care (skills subscale), but only in cancer survivors. All measures of barriers in access to care were directly inter-associated in both groups, but reports of such barriers by cancer survivors showed significant positive associations with scores on anxiety, perceived stress, and post-traumatic symptoms. Interestingly, the perceived social support scores in cancer survivors revealed a positive relationship with depression, anxiety, post-traumatic symptoms, distress, and perceived stress. In summary, the psychosocial variables exhibited stronger associations among them in cancer survivors than in non-cancer participants.

**Figure 1.**
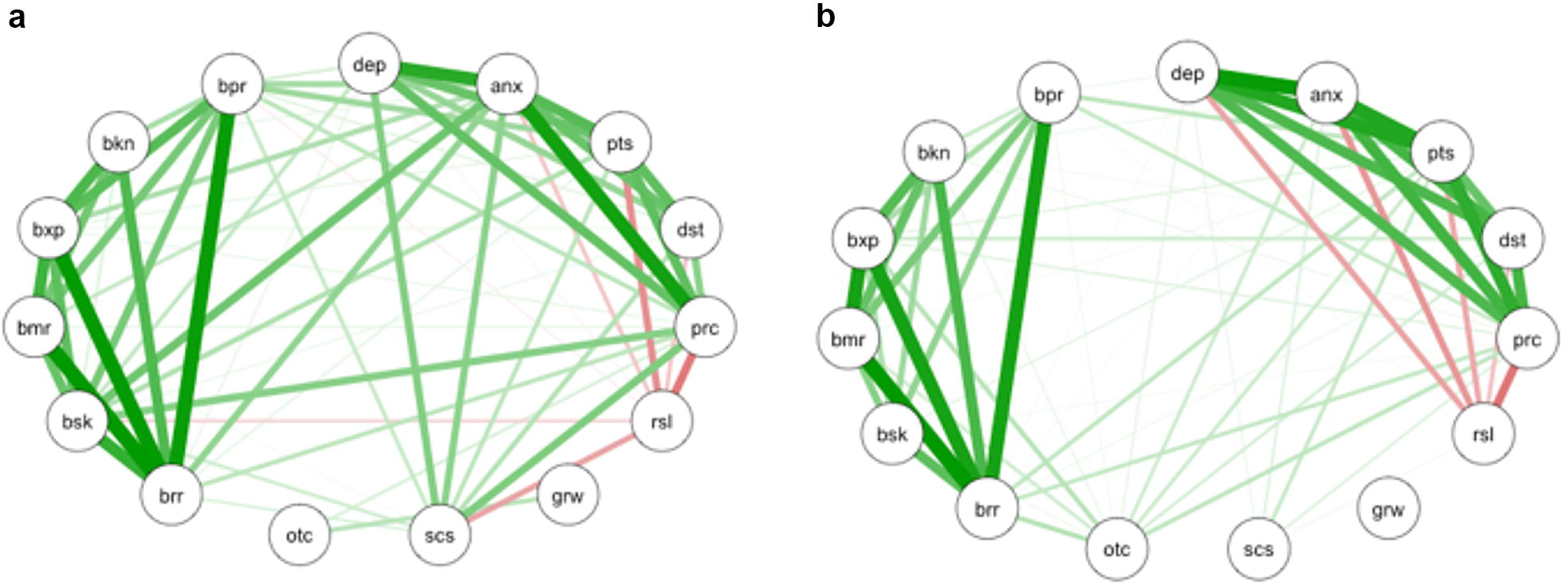
Correlation network of psychosocial assessments in (A) cancer and (B) non-cancer group. Only significant correlations are shown as determined by qgraph. Green lines: positive correlations; red lines: negative correlations. Line thickness shows the strength of the correlation. All comparisons shown are statistically significant. *P* < 0.05. *brr*: barriers to care – total; *bsk*: barriers to care – skills; *bmr*: barriers to care – marginalization; *bxp*: barriers to care – expectations; *bkn*: barriers to care – knowledge and beliefs; *bpr*: barriers to care – pragmatics; *dep*: depressive symptomatology; *anx*: anxiety symptomatology; *pts*: post-traumatic stress disorder symptoms; *dst*: distress; *prc*: perceived stress; *rsl*: resilience; *grw*: post-traumatic growth; *scs*: perceived social support; *otc*: natural disaster outcomes.

Linear regression analyses confirmed a significant association between barriers in access to care and cancer status (Supplementary Table 1; *β* = 7.28, 95% CI: 0.58-13.97, *p*=0.03). Being a cancer survivor increased the barriers to access to care score by 7.28 points. However, after controlling for age and time of recruitment after the hurricane (*β* = 7.01, 95% CI: −0.59-14.08, *p*=0.06) the model failed to show a significant association between barriers in access to care and cancer status (Supplemental Table 1). Only when we adjusted our model to account for recruitment time after the hurricane, the association became significant. Specifically, being a cancer survivor increased barriers in access to care score by 7.38 after controlling for time of recruitment (Supplemental Table 1; *β* = 7.38, 95% CI: 0.75-14.00, *p*=0.03). This model revealed that cancer status and time of recruitment explained 7.4% of the scores in barriers in access to care. The regression formula was: Barriers score = 23.04 + 7.38*cancer status – 1.47*recruitment time. The predicted score of barriers in access to care is 28.85 when an individual has cancer and if he or she was recruited in the month following the hurricane.

### Serum Cytokine Analyses

Serum was obtained from 40 cancer survivors and 35 non-cancer participants. Figure 2a shows a heatmap visualization of cytokine expression by group. Figure 2b-c shows cytokines that were significantly upregulated in the cancer survivor group when compared to non-cancer participants. The most significantly upregulated cytokines were CD31, BDNF, TFF3, Serpin E-1, Vitamin D BP, VCAM-1, Osteopontin, Chitinase 3 like 1, MMP-9 and MIF (Figure 2c). To determine possible protein-protein association networks we data-mined the STRING V11 (22). Figure 2d depicts molecular relationships, interactions, and pathway associations between cytokines that were significantly upregulated in cancer survivors compared to non-cancer participants. To understand biological processes that could be modulated by these cytokines, we performed pathway enrichment analyses using STRING v11. Table 2 depicts KEGG pathway enrichment data (23-25) based on significantly upregulated cytokines identified in cancer survivors compared to non-cancer participants. These analyses revealed significantly enriched pathways that included leukocyte migration, PI3K-AKT and TNF signaling pathways, cell and focal adhesion, Ras and MAPK signaling, among other pathways (Supplemental Table 2).

**Figure 2.**
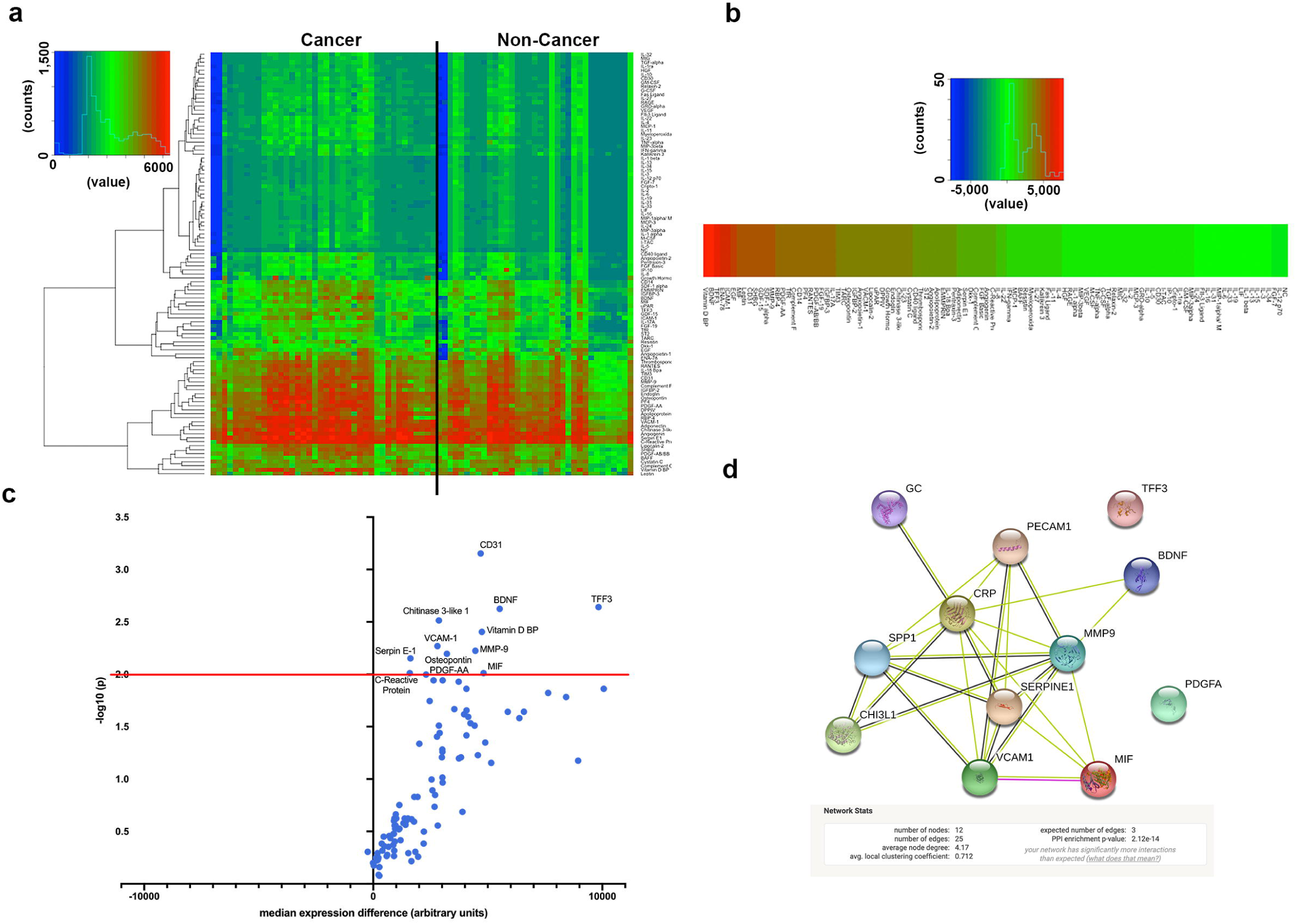
Serum cytokine expression in cancer and non-cancer participants. (A) Heatmap depicting cytokine expression among cancer and non-cancer participants. (B) Individual cytokine differences between cancer and non-cancer participants. Higher (red) to lower (green) differences among groups. (C) Volcano plot depicting cytokine changes (x-axis) and p-values (statistical significance was established as *p* < 0.01; y-axis). (D) String diagram depicting molecular relationships, interactions, and pathway associations between significantly upregulated cytokines.

Furthermore, we explored if there was a correlation between the top ten significantly upregulated cytokines and psychosocial measures using the Spearman’s Rho correlation procedure. Figure 3 illustrates a visual representation of a correlation network with significant associations done using corrplot in RStudio 1.1.463. First, we evaluated the whole cohort and found that most cytokines had a significant positive correlation with perceived social support (Figure 3a and Supplemental Figure 1a). When the cohort was divided into groups our data show that this effect was only observed in the non-cancer group (Figure 3b-c and Supplemental Figure 1b-c). Interestingly, in the cancer survivor group, BDNF, MMP9 and Osteopontin, all known to promote disease progression (26-28), had significant positive correlations with several measures related to barriers in access to care (Figure 3c and Supplemental Figure 1c).

**Figure 3.**
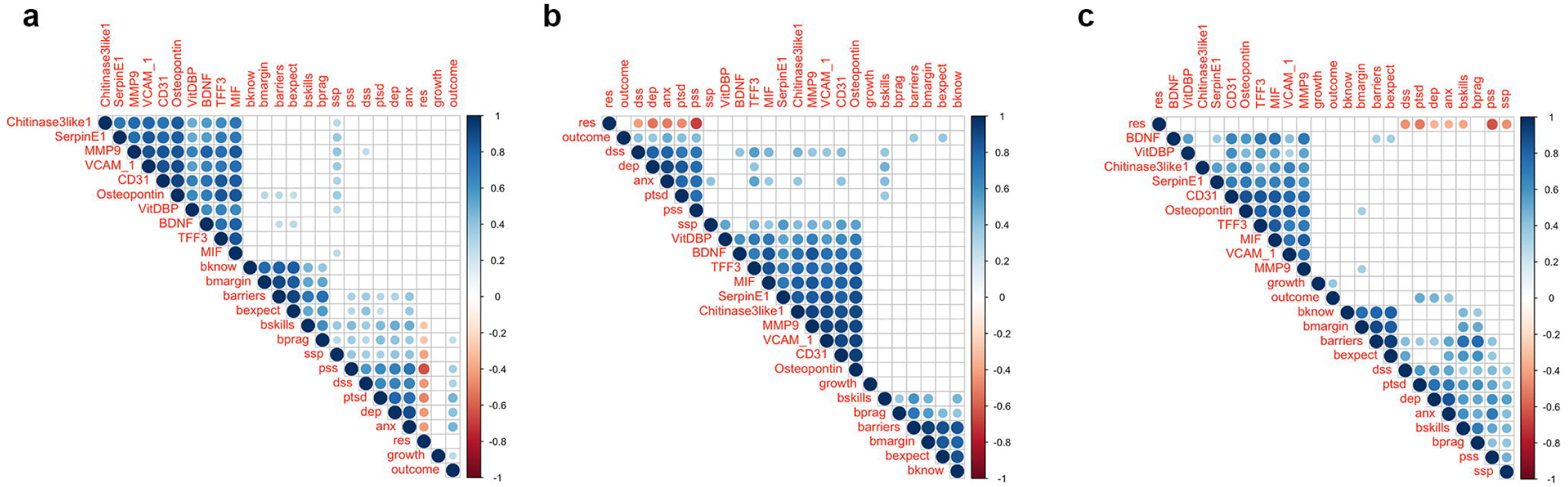
Correlation of top significantly expressed cytokines with psychosocial measurements. Top differentially expressed cytokines (by *P*-value) were subjected to Spearman correlation analyses to identify associations with psychosocial measurements among (A) whole cohort, (B) non-cancer participants and (C) cancer group. White squares = no significant association; Blue circles = significant positive association; Red = significant negative associations. Color intensity reflects stronger associations as determined by the correlation coefficient value (Y-axis). *P* < 0.05. *bknow*: barriers to care – knowledge and belief; *bmargin*: barriers to care – marginalization; *barriers*: barriers to care - total; *bexpect*: barriers to care – expectations; *bskills*: barriers to care – skills; *bprag*: barriers to care: pragmatics; *ssp*: social support; *pss*: perceived stress; *dss*: distress; *ptsd*: post-traumatic stress syndrome symptoms; *dep*: depressive symptomatology; *anx*: anxiety symptoms; *res*: resilience; *growth*: post-traumatic growth; *outcome*: natural disaster outcomes.

## Discussion

Our data show that cancer survivors that faced HM and its aftermath suffered from lack of access to care leading to increased health disparities among Puerto Rican cancer patients. In fact, not only were cancer survivors presented with more barriers in accessing medical care, but our research also revealed direct relationships with anxiety, perceived stress, and post-traumatic symptomatology that were only observed in cancer survivors. Linear regression analysis confirmed that being a cancer survivor predicted more barriers to receiving health care along with closeness to the events related to a natural disaster (such as HM). Moreover, several inflammatory cytokines were found to be significantly upregulated in cancer survivors while pathway enrichment analyses showed that these were associated with activation of tumor-promoting pathways such as those mediated by MAPK, PI3K-AKT, Ras and TNF. Further, we uncovered a positive correlation between several cytokines and perceived social support in cancer survivors. Also, in cancer survivors BDNF, MMP9 and Osteopontin (all associated to pro-tumoral processes) levels were associated with lack of access to care.

As expected, the effects of HM in PR infrastructure affected cancer survivors as well as non-cancer participants. In general, participant’s responses in most of the psychosocial measures suggest that both groups were affected equally. Although psychological distress symptoms in the total sample showed a negative and significant relationship with resilience, group analysis revealed that the association of resilience with symptomatology of depression in the group of cancer survivors was not significant. These findings show that compared to non-cancer participants, cancer survivors report greater severity in the symptoms of depression and greater resiliency. This observation is unexpected, but it can be explained by patients’ tendency to conceal emotional distress to protect loved ones from worrying. Also, in cancer survivors, social support was positively correlated with distress. This association could be explained by Hispanic cultural values that emphasize supportive family relationships where families are prioritized before individual’s well-being. On the other hand, the survivor’s resiliency was negatively related to barriers in access to care. Exacerbations in such barriers after the hurricane for a prolonged time can put them at risk of worsening prognoses, both physically and psychologically.

Our data identified several cytokines that have been associated with inflammatory processes, biobehavioral factors and cancer biology (6, 8, 29). For example, CD31 has been associated to angiogenic processes and cancer progression that have been shown to be promoted by chronic stress in preclinical models of cancer (6, 30). Moreover, BDNF was recently shown to be involved in cancer progression and to be modulated by chronic stress and activation of the SNS (26). Our data also identified several signaling pathways that were enriched by cytokines found to be significantly induced. These included PI3K-Akt (31, 32), MAPK (6), Ras and TNF (6, 8); signaling nodes that are well-known to play key roles in cancer biology. Finally, induction of BDNF and MMP9 was associated with barriers to access to care, in itself a potential source of distress. This is an important observation as these cytokines have been reported to play key roles in biobehavioral effects on cancer (26, 27). They have also been associated with metastases, cancer progression and activation of the SNS (26, 27). These data suggest that psychosocial outcomes in the aftermath of a natural disaster could potentially play a role in cancer biology by promoting tumor-associated processes.

### Study Limitations

Even though our findings provide new light of how a natural disaster can affect cancer survivors, we acknowledge that our results are limited by not controlling for type of cancer, or between subjects with active disease vs. survivors, cancer stage, age, sex, and other comorbidities. Our analyses were also limited by not knowing the previous psychological history of participants. We acknowledge that the psychometric properties of our newly developed Natural Disaster Outcomes Questionnaire needs further evaluation. Our cohort did not include a cancer group that was not exposed to a natural disaster, so we cannot conclude that the biological differences seen were due to a cancer diagnosis and/or the hurricane.

### Clinical Implications of the Study

We consider that cancer survivors can benefit from positive or appropriate social support that in the case of patients can help reduce negative psychosocial comorbidities and increase medical adherence. Several studies have highlighted the importance of social support networks and how this support can help overcome a number of common barriers to treatment, leading to better adjustment to the cancer diagnosis (33, 34). We propose that it is of paramount importance to identify factors that promote and influence resilience and well-being among Hispanic/Latinos facing a chronic and/or terminal illness following natural disasters. Communities and relevant groups could help by pinpointing other factors that may exacerbate cancer survivors perceived stress after a natural disaster.

### Conclusions

Our findings support changes in public policy that includes plans to ensure prompt access to treatment and specialists for cancer survivors and mitigate any barriers to care in the aftermath of a hurricane. These processes will lead to better care and promote population well-being in the face of natural disasters as it is essential for stakeholders to consider the psychological needs of cancer survivors.

## Data Availability

The data that support the findings of this study are available from the corresponding author upon reasonable request.

## Acknowledgements

We would like to acknowledge support from Hospital San Lucas, the PHSU/MCC U54 Partnership’s PRBB staff, Drs. Zuleika Diaz and Hector Velez, Rayo de Luz y Esperanza and Esfuerzate y Se Valiente support groups.

## Author Contributions

MR-R, IF, HJ, EMC, GNA-P analyzed data, performed statistical analyses and wrote the manuscript. RH, ZR, NT, ET, AM collected psychosocial data, biological samples and analyzed data. DA, LM, CBC-E and JP-M analyzed biological samples and performed statistical analyses. All authors read and approved the final manuscript.

